# Protection against Omicron re-infection conferred by prior heterologous SARS-CoV-2 infection, with and without mRNA vaccination

**DOI:** 10.1101/2022.04.29.22274455

**Authors:** Sara Carazo, Danuta M. Skowronski, Marc Brisson, Chantal Sauvageau, Nicholas Brousseau, Rodica Gilca, Manale Ouakki, Sapha Barkati, Judith Fafard, Denis Talbot, Vladimir Gilca, Geneviève Deceuninck, Christophe Garenc, Alex Carignan, Philippe De Wals, Gaston De Serres

## Abstract

**Importance:** Omicron is phylogenetically- and antigenically-distinct from earlier SARS-CoV-2 variants and the original vaccine strain. Protection conferred by prior SARS-CoV-2 infection against Omicron re-infection, and the added value of vaccination, require quantification.

**Objective:** To estimate protection against Omicron re-infection and hospitalization conferred by prior heterologous SARS-CoV-2 (non-Omicron) infection and/or up to three doses of (ancestral, Wuhan-like) mRNA vaccine.

**Design:** Test-negative study between December 26 (epi-week 52), 2021 and March 12 (epi-week 10), 2022.

**Setting:** Population-based, province of Quebec, Canada

**Participants:** Community-dwelling ≥12-year-olds tested for SARS-CoV-2.

**Exposures:** Prior laboratory-confirmed infection with/without mRNA vaccination.

**Outcomes:** Laboratory-confirmed SARS-CoV-2 re-infection and hospitalization, presumed Omicron by genomic surveillance. The odds of prior non-Omicron infection with/without vaccination were compared among Omicron cases/hospitalizations versus test-negative controls (single randomly-selected per individual). Adjusted odds ratios controlled for age, sex, testing-indication and epi-week. Analyses were stratified by severity and time since last non-Omicron infection or vaccine dose.

**Results:** Without vaccination, prior non-Omicron infection reduced the Omicron re-infection risk by 44% (95%CI:38-48), decreasing from 66% (95%CI:57-73) at 3-5 months to 35% (95%CI:21-47) at 9-11 months post-infection and <30% thereafter. The more severe the prior infection, the greater the risk reduction: 8% (95%CI:17-28), 43% (95%CI:37-49) and 68% (95%CI:51-80) for prior asymptomatic, symptomatic ambulatory or hospitalized infections. mRNA vaccine effectiveness against Omicron infection was consistently significantly higher among previously-infected vs. non-infected individuals at 65% (95%CI:63-67) vs. 20% (95%CI:16-24) for one-dose; 68% (95%CI:67-70) vs. 42% (95%CI:41-44) for two doses; and 83% (95%CI:81-84) vs. 73% (95%CI:72-73) for three doses.

Infection-induced protection against Omicron hospitalization was 81% (95%CI: 66-89) increasing to 86% (95%CI:77-99) with one, 94% (95%CI:91-96) with two and 97%(95%CI:94-99) with three mRNA vaccine doses. Two-dose effectiveness against hospitalization among previously-infected individuals did not wane across 11 months and did not significantly differ from three-dose effectiveness despite longer follow-up (median 158 and 27 days, respectively).

**Conclusions and relevance:** Prior heterologous SARS-CoV-2 infection provided substantial and sustained protection against Omicron hospitalization, greatest among those also vaccinated. In the context of program goals to prevent severe outcomes and preserve healthcare system capacity, >2 doses of ancestral Wuhan-like vaccine may be of marginal incremental value to previously-infected individuals.

## Introduction

The Omicron lineage of SARS-CoV-2 emerged in November 2021 as a phylogenetically and antigenically-distinct variant of concern (VOC). With its greater intrinsic transmissibility and capacity for immunological escape, Omicron spread in an unprecedented way, causing the most intense epidemic surge since the beginning of the COVID-19 pandemic, even among countries with high vaccine coverage.^1–6^

The extent to which prior infection and/or vaccination may have modulated risk during the substantial Omicron wave requires better understanding. In this population-based analysis, we quantified protection conferred by prior heterologous infection against Omicron re-infection and hospitalization, stratified by timing and severity of prior infection. We further assessed the effectiveness of up to three doses of heterologous mRNA (ancestral Wuhan-like) vaccine against Omicron among previously SARS-CoV-2-infected versus non-infected individuals.

## Methods

### Study design

Test-negative design was conducted among community-dwelling residents ≥12-years-old in the province of Quebec, Canada with specimen collection between December 26 (epi-week 52), 2021 and March 12 (epi-week 10), 2022 tested for SARS-CoV-2 by nucleic acid amplification testing (NAAT).

Cases were SARS-CoV-2 test-positive during the study period. Upon first being identified as a case during the study period, individuals were censored from further case or control contribution. Controls were SARS-CoV-2 test-negative during the study period; for individuals with multiple negative tests, a single negative specimen per individual was randomly selected.

The odds of prior SARS-CoV-2 infection before the study period, with/without vaccination, were compared between specimens identified to be cases (or hospitalizations) versus controls during the study period. The same set of controls was used to assess protection against infection and hospitalization.

### Data sources

In Quebec, NAAT was limited to prioritized groups and those with severe outcomes during the first pandemic wave but from July 2020 to January 2022 became broadly accessible to the general population through community-based assessment centres. From January 5, 2022, NAAT was restricted due to limited laboratory capacity and availability of rapid antigen detection testing (RADT) during the substantial Omicron wave. It was available to patients consulting or admitted to hospital; to healthcare workers (HCWs) and their families; and to residents in closed settings, but was not routinely-available to asymptomatic contacts or mild cases not requiring medical consultation.^7^

Specimens were sampled from the provincial laboratory database which includes all NAAT for SARS-CoV-2 conducted in Quebec since pandemic start. The database also captures testing indication, including categories of: (1) symptomatic, tested outside medical care setting (e.g. assessment centres); (2) symptomatic, during emergency room visit or hospitalization; (3) symptomatic, among HCWs; (4) asymptomatic, during outbreaks in care facilities/closed settings; (5) asymptomatic, screening at hospital pre-admission; (6) asymptomatic contacts; (7) other asymptomatic; and (8) any other reason, combined.

Specimens within the provincial laboratory database were linked through unique personal identifying numbers (PINs) to the: a) provincial immunization registry that specifies vaccine status for all Quebec residents including type, dates and number of vaccine doses; b) database of all COVID-19 cases reported to public health including demographic/clinical details; c) administrative hospitalization database; and d) VOC screening and/or whole genome sequencing results.

### Outcome and primary infection definitions

The main outcome was any NAAT-confirmed SARS-CoV-2 infection during the study period.^8^ COVID-19 hospitalization, defined by admission ≥24-hours and within 14 days of a SARS-CoV-2 positive specimen, was also assessed.

Reinfection was defined as the first SARS-CoV-2 positive result during the study period identified ≥90 days after a primary infection.^9^ Primary infection was defined by the first positive specimen from the beginning of the pandemic to ≥90 days before specimen collection during the study period. Those without such record in the laboratory database were considered previously uninfected (or infection-naïve). Those for whom SARS-CoV-2 reinfection was previously documented before December 26, 2021 and all specimens collected <90 days after a first positive specimen were excluded.

### VOC attribution

VOC detection provincially varied in response to changing epidemic patterns, case load and laboratory capacity as well as the profile of emerging and identified VOC, as described in **Supplementary Figure 1**.^8^ For test-positive specimens collected between February 1 and October 8, 2021, Alpha, Beta, Gamma or Delta VOC-status was based on individual-level genetic characterization undertaken on all SARS-CoV-2 detections provincially. VOC status was otherwise presumptive based on provincial-level genomic surveillance. Before February 1, 2021, primary infections were designated “pre-VOC”. From October 9 to December 12, 2021, which spans to the last day of specimen collection meeting the ≥90-day spacing required by the re-infection definition, primary infections were designated Delta. During the study period, spanning December 26, 2021 to March 12, 2022, all case viruses were assumed to be due to Omicron.

### Vaccination definitions

The publicly-funded COVID-19 immunization campaign began in Quebec on December 14, 2020. Community-based vaccination sequentially prioritized the oldest to the youngest down to 5-years of age. Second doses were deferred up to 16 weeks with third doses also beginning with the oldest in December 2021 (**Supplementary Figure 1**).^10,11^

Only the effects of mRNA vaccination were assessed. Single-dose mRNA vaccination was defined by receipt of one dose of BNT162b2 (Pfizer-BioNTech) or mRNA-1273 (Moderna) vaccines ≥21 days before specimen collection. Two-dose and three-dose vaccination was defined by receipt ≥7 days before specimen collection. To exclude immunocompromised persons for whom a 4-week interval between 2^nd^ and 3^rd^ doses was recommended,^12^ only participants receiving the 3^rd^ dose ≥90 days after the 2^nd^ dose were included, as per more-routine recommendation in Quebec.^13^

### Exclusion criteria

Additional exclusions include specimens collected from participants for whom: the reason for testing was the confirmation of COVID-19 recovery or confirmation of a positive RADT; vaccination date or interval between doses was invalid; first dose was received <21 days or second or third doses <7 days before testing; or a non-mRNA vaccine (Astra-Zeneca or Janssen) was received. Hospitalization analysis further excluded those whose testing indication was for pre-admission screening.

### Statistical analyses

Exposure was analysed as a combination of prior primary infection (PI) and vaccine (V) receipt as follows: PI non-vaccinated (PI-NV); PI before first, second or third vaccine dose (PI-V1, PI-V2, PI-V3); PI after first, second, or third vaccine dose (V1-PI, V2-PI, V3-PI); PI after first but before second vaccine dose (V1-PI-V2); PI after first but before second and third vaccine doses (V1-PI-V3); PI after second but before third vaccine doses (V2-PI-V3). Those with no infection (NI) previously were also sub-categorized based upon the number of vaccine doses received as NI-NV, NI-V1, NI-V2, and NI-V3 (**Supplementary Figure 2**).

The main comparator for all analyses was the NI-NV group. The odds of having experienced PI with/without vaccination (PI-NV, PI-V1, PI-V2, PI-V3) or of NI with vaccination (NI-V1, NI-V2, NI-V3) were compared among Omicron cases/hospitalizations versus test-negative controls during the study period through adjusted odds ratios (aOR) controlling for age, sex, testing-indication and epi-week. Effectiveness/protection was derived as 1-aOR with 95% confidence intervals (CI).

Analyses were stratified by age group, PI severity and variant status, time since PI, and time since last event (PI or vaccine dose). Participants with prior infection (PI-NV, PI-V1, PI-V2, PI-V3) were also directly compared to vaccinated individuals with no infection previously (NI-V1, NI-V2, NI-V3).

Analyses were performed using SAS version 9.4 (SAS Institute Inc).

### Ethics statement

The study was conducted under the legal mandate of the National Director of Public Health of Quebec under the Public Health Act and was also approved by the Research Ethics Board of the Centre hospitalier universitaire de Québec-Université Laval.

## Results

### Study population

Among 1,778,623 specimens tested by NAAT overall during the study period, data linkage was successful for 1,754,358 (98.6%). After applying eligibility criteria and exclusions detailed in **Supplementary Figure 3**, and randomly selecting one negative specimen per individual as control, 224,007 test-positive cases and 472,432 test-negative controls were included in analyses.

### Baseline characteristics

The characteristics of cases and controls by prior PI status are displayed in **Table 1**.

**Table 1.** Characteristics of Omicron cases and controls stratified by primary SARS-CoV-2 (non-Omicron) infection history

|  | Cases (Omicron) |  | Controls |  |
| --- | --- | --- | --- | --- |
|  | Prior primary infection (PI: non-Omicron) | No infection (NI) previously | Prior primary infection (PI: non-Omicron) | No infection (NI) previously |
| **Total N** | 9505 | 214502 | 29712 | 442720 |
|  | **n (%)** | **n (%)** | **n (%)** | **n (%)** |
| **Female sex** | 6681 (70.3) | 132726 (61.9) | 20668 (69.6) | 281290 (63.5) |
| **Age (years)** |  |  |  |  |
| 12-17 | 321 (3.4) | 11940 (5.6) | 886 (3.0) | 13756 (3.1) |
| 18-49 | 7120 (74.9) | 137679 (64.2) | 18845 (63.4) | 211324 (47.7) |
| 50-69 | 1789 (18.8) | 49302 (23.0) | 7668 (25.8) | 119015 (26.9) |
| ≥70 | 275 (2.9) | 15581 (7.3) | 2313 (7.8) | 98625 (22.3) |
| **Healthcare worker** | 3821 (40.2) | 45251 (21.1) | 12952 (43.6) | 98571 (22.3) |
| **Year-epi-week of specimen collection (calendar start/end date)** |  |  |  |  |
| 2021-52 (Dec 26, 2021 – Jan 1, 2022) | 2599 (27.3) | 70753 (33.0) | 6109 (20.6) | 90962 (20.6) |
| 2022-01 (Jan 2 – 8, 2022) | 2460 (25.9) | 54928 (25.6) | 5255 (17.7) | 71372 (16.1) |
| 2022-02 to 2022-04 (Jan 9 – 29, 2022) | 2583 (27.2) | 46624 (21.7) | 8735 (29.4) | 117801 (26.6) |
| 2022-05 to 2022-07 (Jan 30 – Feb 19, 2022) | 1258 (13.2) | 27980 (13.0) | 5763 (19.4) | 91917 (20.8) |
| 2022-08 to 2022-10 (Feb 20 – Mar 12, 2022) | 605 (6.4) | 14217 (6.6) | 3850 (13.0) | 70668 (16.0) |
| **Indication for testing** |  |  |  |  |
| Symptomatic, outside care setting | 3233 (34.0) | 102260 (47.7) | 4050 (13.6) | 74839 (16.9) |
| Symptomatic, emergency room | 284 (3.0) | 12551 (5.9) | 2000 (6.7) | 46267 (10.5) |
| Symptomatic, healthcare worker | 3262 (34.3) | 49111 (22.9) | 4117 (13.9) | 46454 (10.5) |
| Asymptomatic, closed setting outbreak | 1009 (10.6) | 13935 (6.5) | 7776 (26.2) | 88078 (19.9) |
| Asymptomatic, hospital pre-admission | 270 (2.8) | 8157 (3.8) | 4450 (15.0) | 102848 (23.2) |
| Asymptomatic contact | 687 (7.2) | 17095 (8.0) | 2608 (8.8) | 39056 (8.8) |
| Asymptomatic other | 505 (5.3) | 5406 (2.5) | 3396 (11.4) | 28075 (6.3) |
| Other reasons combined | 255 (2.7) | 5987 (2.8) | 1315 (4.4) | 17103 (3.9) |
| **PI VOC status** |  |  |  |  |
| Non-VOC | 7424 (78.1) | NA | 22826 (76.8) | NA |
| Alpha | 613 (6.5) | NA | 1955 (6.6) | NA |
| Delta | 253 (2.7) | NA | 1458 (4.9) | NA |
| Other / unknown<sup>a</sup> | 1215 (12.8) | NA | 3427 (11.7) | NA |
| **Interval between PI and specimen collection** |  |  |  |  |
| 90-182 days (3-5 months) | 372 (3.9) | NA | 1791 (6.0) | NA |
| 183-274 days (6-8 months) | 590 (6.2) | NA | 1687 (5.7) | NA |
| 275-364 days (9-11 months) | 1904 (20.0) | NA | 4931 (16.6) | NA |
| 365-547 days (12-18 months) | 4747 (49.9) | NA | 14355 (48.3) | NA |
| 548-730 days (19-24 months) | 1892 (19.9) | NA | 6948 (23.4) | NA |
| Median (interquartile range), days | 407 (354 – 480) | NA | 414 (356 – 514) | NA |
| Range, days | 90 – 714 | NA | 90 - 731 | NA |
| **Severity of PI** |  |  |  |  |
| Asymptomatic | 1183 (12.5) | NA | 2685 (9.0) | NA |
| Symptomatic non-hospitalized | 8123 (85.5) | NA | 25615 (86.2) | NA |
| Symptomatic hospitalized | 199 (2.1) | NA | 1412 (4.8) | NA |

<sup>a</sup> PIs during periods with mixed VOC circulation without individual-level genotyping or identified to be Beta (n=2) or Gamma (n=5) VOCs
Abbreviations: NA, non-applicable; NI, no infection previously; PI, prior primary infection; VOC, variant of concern

Among all 224,007 cases during the study period, 9,505 (4.2%) were reinfections (**Table 2**). With respect to vaccination status, 17,633 (7.9%) cases overall were NI-NV, but most (142,326; 63.5%) were NI-V2. Conversely, 915 (0.4%) were PI-NV, 347 (0.2%) received any number of vaccine doses before their PI and 8243 (3.7%) after their PI. Among the 5,057 COVID-19 hospitalizations during the study period, 64 (1.3%) were cases with prior PI; whereas, no deaths were identified among cases with prior PI.

**Table 2.** Omicron cases and controls stratified by outcome severity and primary SARS-CoV-2 (non-Omicron) infection and vaccination history

|  | Cases (Omicron) |  |  | Controls | All |
| --- | --- | --- | --- | --- | --- |
|  | Infection | Hospitalization<sup>a</sup> | Death<sup>a</sup> |  |  |
|  | N (%) | N (%) | N (%) | N (%) | N (%) |
| **Overall** | **224007** | **5057** | **920** | **472432** | **696439** |
| **Prior primary SARS-CoV-2 (non-Omicron) infection (PI)** | **9505 (4.2)** | **64 (1.3)** | **0 (0.0)** | **29712 (6.3)** | **39217 (5.6)** |
| PI-NV | 915 (0.4) | 13 (0.3) | 0 (0.0) | 1817 (0.4) | 2732 (0.4) |
| **Vaccination after PI:** | **8243 (3.7)** | **48 (0.9)** | **0 (0.0)** | **26006 (5.5)** | **34249 (4.9)** |
| PI-V1 | 2102 (0.9) | 18 (0.4) | 0 (0.0) | 4906 (1.0) | 7008 (1.0) |
| PI-V2 | 5038 (2.3) | 23 (0.5) | 0 (0.0) | 13942 (3.0) | 18980 (2.7) |
| PI-V3 | 1103 (0.5) | 7 (0.1) | 0 (0.0) | 7158 (0.2) | 8261 (1.2) |
| **Vaccination before PI:** | **347 (0.2)** | **3 (0.1)** | **0 (0.0)** | **1889 (0.4)** | **2236 (0.3)** |
| V1-PI | 13 (0.0) | 0 (0.0) | 0 (0.0) | 54 (0.0) | 67 (0.0) |
| V2-PI | 107 (0.1) | 1 (0.0) | 0 (0.0) | 474 (0.1) | 581 (0.1) |
| V3-PI | 2 (0.0) | 0 (0.0) | 0 (0.0) | 5 (0.0) | 7 (0.0) |
| V1-PI-V2 | 123 (0.1) | 1 (0.0) | 0 (0.0) | 385 (0.1) | 508 (0.1) |
| V1-PI-V3 | 66 (0.0) | 1 (0.0) | 0 (0.0) | 485 (0.1) | 551 (0.1) |
| V1-PI-V3 | 36 (0.0) | 0 (0.0) | 0 (0.0) | 486 (0.1) | 522 (0.1) |
| **No infection (NI) with SARS-CoV-2 previously** | **214502 (95.8)** | **4993 (98.7)** | **920 (100)** | **442720 (93.7)** | **657222 (94.4)** |
| NI-NV | 17633 (7.9) | 1399 (27.7) | 240 (26.1) | 20997 (4.4) | 38630 (5.6) |
| NI-1d | 3899 (1.7) | 146 (2.9) | 32 (3.5) | 5307 (1.1) | 9206 (1.3) |
| NI-2d | 142326 (63.5) | 2140 (42.3) | 377 (41.0) | 185780 (39.3) | 328106 (47.1) |
| NI-3d | 50644 (22.6) | 1308 (25.9) | 271 (29.5) | 230636 (48.8) | 281280 (40.4) |

Abbreviations: NI, no infection previously; NI-NV, no infection previously non-vaccinated; NI-V1, no infection previously, one vaccine dose; NI-V2, no infection previously, two vaccine doses; NI-V3, no infection previously, three vaccine doses; PI, prior primary infection; PI-NV, prior infection non-vaccinated; PI-V1, prior infection before one vaccine dose; PI-V2, prior infection before two vaccine doses; PI-V3, prior infection before three vaccine doses; V1-PI, prior infection after one vaccine dose; V1-PI-V2, prior infection after first but before second vaccine dose; V1-PI-V3, prior infection after first but before second and third vaccine doses; V2-PI, prior infection after two vaccine doses; V2-PI-V3, prior infection after second but before third vaccine dose; V3-PI, prior infection after three vaccine doses
<sup>a</sup>Hospitalizations (1749) and deaths (139) for cases who had "preadmission to health facility" as indication for testing were excluded

Among all 472,432 controls during the study period, PI was identified among 29,712 (6.3%) (**Table 2**). With respect to vaccination status, 20,997 (4.4%) controls overall were NI-NV but most (230,636; 48.8%) were NI-V3. Conversely, 1,817 (0.4%) were PI-NV, 1,889 (0.4%) received any number of vaccine doses before their PI and 26,006 (5.5%) after their PI.

Among the 9,505 reinfections during the study period, most (78%) had a prior PI genetically-categorized as pre-VOC, likely reflecting a longer period for accrual (notably pre-vaccine roll-out) compared to Alpha, Delta or other/unknown VOC circulation, which instead comprised 7%, 3%, and 12% of PIs, respectively (**Table 1**). The median specimen collection interval between PI and case or control detection was similar (407 vs. 414 days, respectively).

### Prior infection-induced protection against Omicron re-infection, without vaccination

Without vaccination, prior non-Omicron infection reduced the Omicron re-infection risk by 44% (95%CI:38-48) (**Table 3**). The more severe the prior infection, the greater the Omicron risk-reduction: 8% (95%CI:17-28), 43% (95%CI:37-49) and 68% (95%CI:51-80) for prior asymptomatic, symptomatic ambulatory or hospitalized infections, also evident among vaccinated individuals. Protection induced by asymptomatic infection alone was evident for the first 6 months (49%;95%CI:8-72) but not thereafter (**Table 4**).

**Table 3.** Primary SARS-CoV-2 (non-Omicron) infection-induced effectiveness against Omicron re-infection and hospitalization, with/without vaccination (by number of doses), relative to non-vaccinated with no infection history

|  | Omicron Infection |  | Omicron Hospitalization |  |
| --- | --- | --- | --- | --- |
|  | Unadjusted effectiveness (95% CI) | Adjusted effectiveness (95% CI) | Unadjusted effectiveness (95% CI) | Adjusted effectiveness (95% CI) |
| **Prior primary infection** |  |  |  |  |
| Unvaccinated | 40% (35, 45) | 44% (38, 48) | 90% (83, 94) | 81% (66, 89) |
| 1-dose vaccinated | 49% (46, 52) | 65% (63, 67) | 96% (93, 97) | 86% (77, 91) |
| 2-dose vaccinated | 57% (55, 59) | 68% (67, 70) | 98% (97, 99) | 94% (91, 96) |
| 3-dose vaccinated | 82% (80, 83) | 83% (81, 84) | 99% (98, 99) | 97% (94, 99) |
| **No prior infection** |  |  |  |  |
| Unvaccinated | Referent | Referent | Referent | Referent |
| 1-dose vaccinated | 12% (8, 16) | 20% (16, 24) | 60% (53, 67) | 52% (42, 61) |
| 2-dose vaccinated | 9% (7, 11) | 42% (41, 44) | 86% (85, 87) | 76% (74, 78) |
| 3-dose vaccinated | 74% (73, 74) | 73% (72, 73) | 92% (92, 93) | 91% (91, 92) |

<sup>a</sup> Logistic regression models adjusted for age (4 categories), sex, indication for testing and epidemiological week Abbreviations: CI, confidence interval

**Table 4.** Primary SARS-CoV-2 (non-Omicron) infection-induced effectiveness against Omicron re-infection, with/without vaccination (by number of doses) stratified by age, and characteristics of the primary infection, relative to non-vaccinated with no infection history

|  | PI-NV | PI-V1 | PI-V2 | PI-V3 |
| --- | --- | --- | --- | --- |
|  | Adjusted effectiveness<sup>a</sup><br>(95% CI) | Adjusted effectiveness<sup>a</sup><br>(95% CI) | Adjusted effectiveness<sup>a</sup><br>(95% CI) | Adjusted effectiveness<sup>a</sup><br>(95% CI) |
| **Global** | 44% (38, 48) | 65% (63, 67) | 68% (67, 70) | 83% (81, 84) |
| **Age (years)** |  |  |  |  |
| 12-17 | 57% (36, 71) | 78% (70, 83) | 79% (74, 93) | 96% (65, 99) |
| 18-49 | 44% (29, 43) | 62% (60, 65) | 67% (65, 68) | 79% (77, 81) |
| 50-69 | 51% (38, 60) | 71% (66, 75) | 72% (69, 74) | 86% (83, 88) |
| ≥70 | 46% (16, 65) | 79% (65, 87) | 67% (60, 73) | 81% (75, 86) |
| **VOC status of prior PI** |  |  |  |  |
| non-VOC | 29% (20, 37) | 62% (59, 64) | 67% (65, 69) | 81% (79, 82) |
| Alpha | 44% (26, 57) | 73% (68, 78) | 75% (71, 79) | 92% (84, 96) |
| Delta | 67% (57, 75) | 73% (61, 82) | 87% (70, 94) | NE |
| Other / unknown<sup>b</sup> | 50% (38, 60) | 70% (64, 74) | 69% (65, 72) | 86% (81, 90) |
| **Severity of prior PI** |  |  |  |  |
| Asymptomatic | 8% (-17, 28) | 40% (28, 50) | 43% (36, 49) | 66% (59, 73) |
| Symptomatic non-hospitalized | 43% (37, 49) | 66% (64, 68) | 70% (68, 71) | 82% (81, 84) |
| Symptomatic hospitalized | 68% (51, 80) | 79% (67, 86) | 77% (72, 82) | 87% (80, 91) |
| **Interval since PI** |  |  |  |  |
| 3-5 months | 66% (57, 73) | 73% (64, 80) | 88% (76, 94) | NE |
| 6-8 months | 49% (32, 61) | 76% (70, 80) | 78% (74, 81) | 89% (70, 96) |
| 9-11 months | 35% (21, 47) | 65% (61, 69) | 68% (65, 70) | 85% (81, 89) |
| 12-18 months | 29% (17, 38) | 61% (58, 64) | 67% (65, 69) | 80% (78, 82) |
| 19-24 months | 27% (8, 42) | 61% (55, 67) | 67% (64, 70) | 80% (78, 82) |
| **Severity of and interval since PI** |  |  |  |  |
| Asymptomatic <6 months | 49% (8, 72) | 66% (17, 86) | NE | NE |
| Asymptomatic ≥6 months | -5% (-36, 20) | 39% (26, 49) | 42% (35, 49) | 66% (59, 73) |
| Symp non-hosp <6 months | 67% (57, 74) | 73% (63, 81) | 88% (76, 94) | NE |
| Symp non-hosp ≥6 months | 37% (29, 43) | 66% (63, 68) | 69% (68, 71) | 82% (81, 84) |
| Symp hosp <6 months | 81% (52, 92) | 84% (23, 97) | 73% (-162, 97) | NE |
| Symp hosp ≥6 months | 62% (36, 77) | 78% (66, 86) | 77% (72, 82) | 87% (80, 91) |
| **Interval since last vaccination<sup>c</sup>** |  |  |  |  |
| <2 months | NA | 81% (74, 86) | 82% (80, 84) | 83% (81, 84) |
| 2-5 months | NA | 64% (60, 67) | 67% (65, 68) | 80% (76, 84) |
| 6-8 months | NA | 62% (58, 65) | 63% (60, 65) | NE |
| 9-11 months | NA | 61% (54, 67) | 62% (42, 75) | NE |
| 12-14 months | NA | 65% (48, 76) | NE | NE |

<sup>a</sup> Logistic regression models comparing persons with prior primary infection with/without vaccination to those unvaccinated and without prior infection. All estimates adjusted for age (4 categories), sex, indication for testing and epidemiological week.
<sup>b</sup> Cases without genotyping during periods with mixed circulation or cases from beta variant (n=2) or gamma variant (n=5)
<sup>c</sup> Models stratified for delay from last vaccination are not adjusted for epidemiological week due to insufficient number of cases in each strata and high correlation between delay and epidemiological week for those vaccinated with 3 doses.
Abbreviations: CI, confidence interval; hosp, hospitalized; NA, non-applicable; NE, non-estimable; PI, prior primary infection; PI-NV, prior infection non-vaccinated; PI-V1, prior infection before one vaccine dose; PI-V2, prior infection before two vaccine doses; PI-V3, prior infection before three vaccine doses; symp, symptomatic

Prior infection-induced protection varied by VOC status: 29% (95%CI:20-37), 44% (95%CI:26-57) and 67% (95%CI:57-75) for pre-VOC, Alpha and Delta, respectively (**Table 4**). This, however, may also reflect waning over differential time since variant-specific circulation. Prior infection-induced protection against Omicron decreased from 66% (95%CI:57-73) at 3-5 months post-infection, reflecting the more-proximal Delta period, to 49% (95%CI:32-61) at 6-8 months when Alpha foremost contributed, and 35% (95%CI:21-47) at 9-11 months, remaining <30% thereafter and foremost reflecting the more distant pre-VOC period (**Figure 1**).

**Figure 1.**
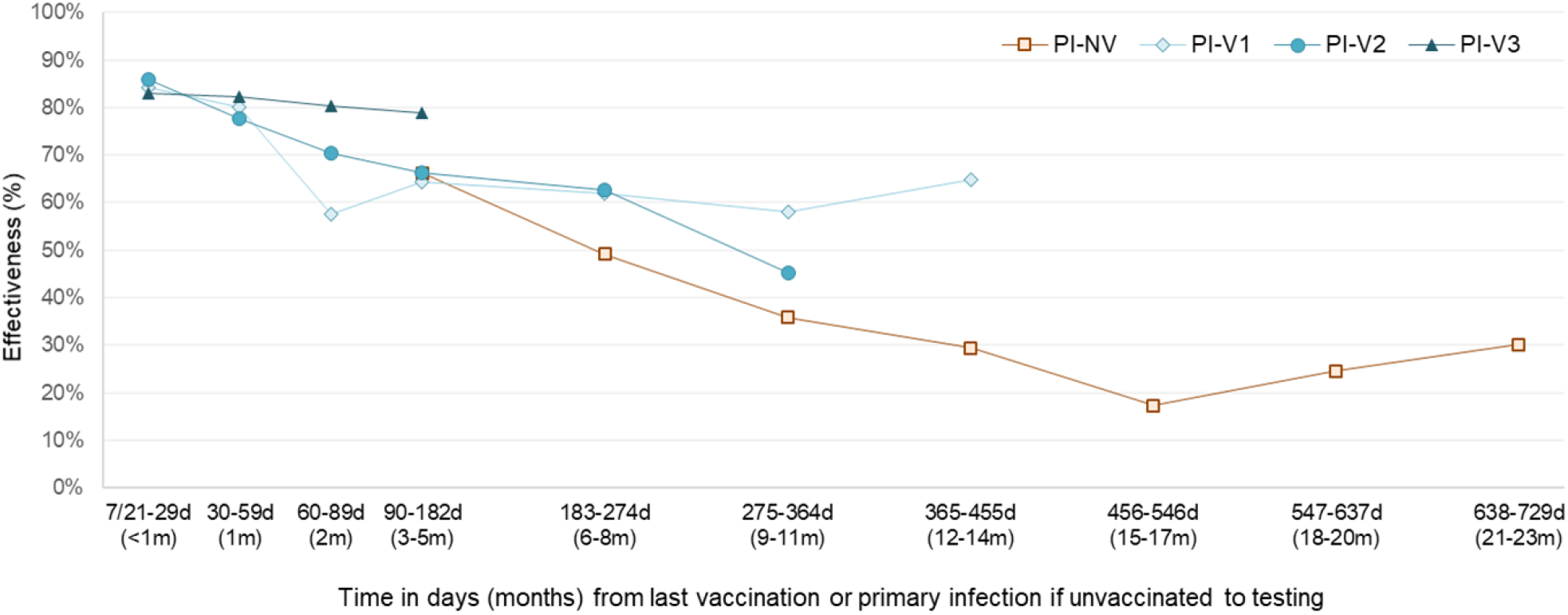
Effectiveness of primary SARS-CoV-2 (non-Omicron) infection against Omicron re-infection, by time since last event (primary infection or vaccination), relative to non-vaccinated with no infection history Abbreviations: d, days; m, months; PI-NV, prior infection non-vaccinated; PI-V1, prior infection before one vaccine dose; PI-V2, prior infection before two vaccine doses; PI-V3, prior infection before three vaccine doses Note: Logistic regression models comparing persons with prior primary infection with/without vaccination to those unvaccinated and without prior infection. All estimates adjusted for age (4 categories), sex, indication for testing and epidemiological week

### Vaccine effectiveness (VE) against Omicron re-infection

VE against Omicron infection was consistently significantly higher among previously-infected vs. non-infected individuals: 65% (95%CI:63-67) vs. 20% (95%CI:16-24) for one dose; 68% (95%CI:67-70) vs. 42% (95%CI:41-44) for two doses; and 83% (95%CI:81-84) vs. 73% (95%CI:72-73) for three doses (**Table 3**). For the same number of vaccine doses, protection against reinfection was similar whether the prior infection came before, between or after vaccination (**Figure 2**).

**Figure 2.**
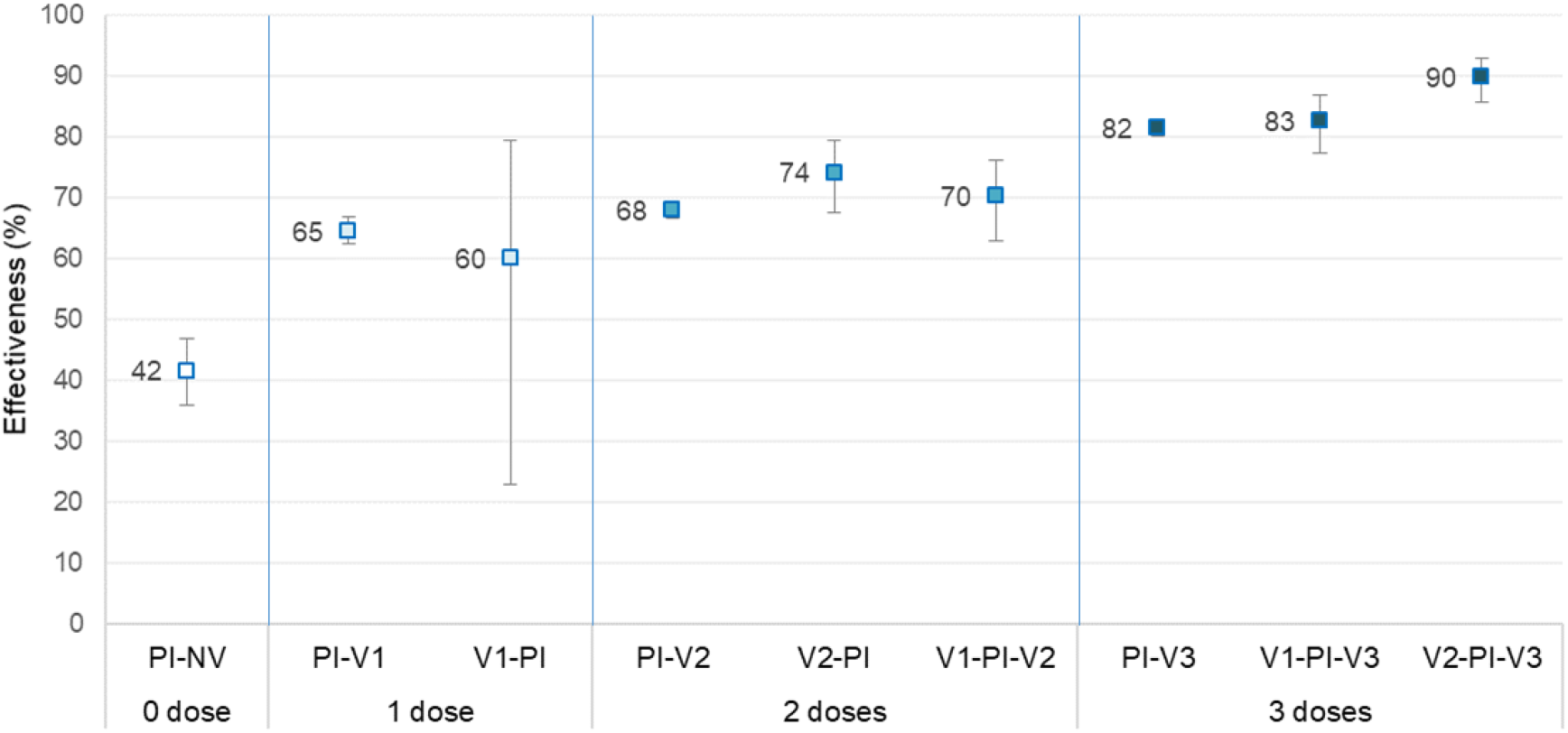
Prior infection and mRNA vaccine effectiveness against Omicron reinfection stratified by number of doses and timing before or after primary SARS-CoV-2 (non-Omicron) infection, relative to non-vaccinated with no infection history Abbreviations: PI-NV, prior infection non-vaccinated; PI-V1, prior infection before one vaccine dose; PI-V2, prior infection before two vaccine doses; PI-V3, prior infection before three vaccine doses; V1-PI, prior infection after one vaccine dose; V1-PI-V2, prior infection after first but before second vaccine dose; V1-PI-V3, prior infection after first but before second and third vaccine doses; V2-PI, prior infection after two vaccine doses; V2-PI-V3, prior infection after second but before third vaccine dose; V3-PI, prior infection after three vaccine doses Note: Logistic regression models comparing persons with prior primary infection with/without vaccination to those unvaccinated and without prior infection. All estimates adjusted for age (4 categories), sex, indication for testing and epidemiological week

Two vaccine doses were significantly less effective than three doses among both previously-infected (68% vs. 83%) and non-infected (42% vs. 73%) individuals, recognizing longer median follow-up time since second vs. third dose among both the previously-infected (158 vs. 27 days) and uninfected (173 vs. 37 days) (not shown). For the same number of doses, the previously-infected were 40-60% better protected against Omicron re-infection than the previously-non-infected (**Supplementary Figure 4)**.

Among the previously-infected, lower VE after one (65%) or two (68%) vs. three doses (83%) may in part be attributed to waning over differential time since vaccination since when standardized for the first 2 months post-vaccination, VE was instead similar at 81%, 82% and 83%, respectively. At 2-5 months post-vaccination, however, corresponding VE was 64%, 67% and 80% and thereafter ranged 60-65% among two-dose recipients (**Table 4, Figure 1**).

### Infection and/or vaccine-induced protection against Omicron hospitalization

Without vaccination, prior non-Omicron infection reduced the Omicron hospitalization risk by 81% (95%CI:66-89) (**Table 3**). Given the high level of infection-induced protection and vaccine coverage, sample size was limited to further stratify by time since or severity of prior infection.

VE against Omicron hospitalization was consistently significantly higher among previously-infected vs non-infected individuals: 86% (95%CI:77-91) vs. 52% (95%CI:42-61) for one dose; 94% (95%CI:91-96) vs.76% (95%CI:74-78) for two doses; and 97% (95%CI:94-99) vs. 91% (95%CI:91-92) for three doses.

Against hospitalization, VE for two doses was similar to three doses among the previously-infected (94% vs. 97%) but was significantly lower among the previously-uninfected (76% vs. 91%), recognizing longer median follow-up time since second vs. third dose, as per above (**Table 3**). For the same number of doses, the previously-infected were significantly 70-80% better protected than the previously non-infected (**Supplementary Figure 5**).

Among previously-infected individuals, two-dose VE against Omicron hospitalization was similar at <6 and 6-11 months post-vaccination (95%; 95%CI:92-97 vs. 93%; 95%CI:86-96); whereas, among the previously non-infected, significant decline in two-dose VE was observed (81%; 95%CI:79-83 vs. 73%; 95%CI:71-75, respectively) (not shown).

## Discussion

We show that a single prior (heterologous, non-Omicron) SARS-CoV-2 infection, without vaccination, reduced the subsequent Omicron re-infection risk by nearly half, including by as much as two-thirds during the first six months and by one-third 9-11 months after primary infection. The more severe the primary infection, the greater the cross-protection against Omicron. Prior infection alone reduced the Omicron hospitalization risk even more substantially by ∼80%, but the best protected were nevertheless those previously-infected and also vaccinated (so-called hybrid protection), among whom two doses reduced the hospitalization risk by 94% with an additional third dose further marginally increasing protection to 97%. Overall, for the same number of vaccine doses, the previously-infected were ∼70-80% better protected against hospitalization than those previously-uninfected. It is reasonable to conclude that absent prior infections and/or vaccination, the population impact of the Omicron surge would have been much worse.

Epidemiological studies conducted during the pre-Omicron period showed prior infection (without vaccination) reduced the risk of reinfection due to the same or different variants by 80-100%, with minimal waning after one year.^14–18^ Few studies have quantified prior infection-induced protection against Omicron, but our findings seem consistent with those available. In Qatar, prior infection was 56% effective against Omicron reinfection and 88% against hospitalization, comparable against re-infection at 3-8 (64%) and ≥15 months (60%) post-infection.^19^ Preprint from the Czech Republic reported decline in infection-induced protection against re-infection from 68% at 2-6 months to 13% >6 months later, whereas protection against hospitalization was 87% and well-maintained.^20^ Overall, strong and sustained protection from prior heterologous infection against severe Omicron outcomes, including the absence of Omicron-associated deaths among survivors of prior infection, appears to be a robust interpretation across the few available studies to date, including our own.^19,20^

A unique contribution of our study is the demonstration that prior asymptomatic infection was also protective, reducing the Omicron re-infection risk by about half during the first six months post-primary infection whereas, thereafter, unvaccinated, asymptomatically-infected individuals had no protective advantage over the infection-naïve. During the pre-Omicron period, a Danish cohort study suggested previously-infected individuals who experienced prior asymptomatic vs. symptomatic infection were at 50% higher risk of Delta reinfection.^21^ To our knowledge, ours is the first study to evaluate such gradient of protection by severity of prior infection against Omicron re-infection.

Based on our findings, prior heterologous SARS-CoV-2 infection conferred protection against Omicron re-infection and hospitalization (44% and 81%, respectively) that was comparable to the effectiveness of two doses of heterologous mRNA vaccine among previously uninfected individuals (42% and 76%, respectively). Similar observations have been reported elsewhere during the pre-Omicron period although these were not fully consistent across studies.^17,22–27^ Elsewhere during the Omicron period, two manuscripts from Qatar instead suggest higher protection conferred by prior infection vs. two-dose vaccination against symptomatic Omicron re-infection;^28,29^ whereas, preprint from the Netherlands reports lower protection from prior infection (25%;95%CI:21-29) vs. two-dose vaccination (33%;95%CI:31-35).^30^ Additional studies may be needed to clarify these comparisons, recognizing the challenge in standardizing based upon the time since last event (infection or vaccination).

Immunogenicity studies conducted during the pre-Omicron period show a single BNT162b2 vaccine dose among previously-infected individuals elicited robust antibody and T cell responses, exceeding two-dose response in the infection-naïve.^31–34^ In epidemiological studies conducted pre-Omicron, one or two BNT162b2 doses in Israel reduced the pre-Omicron re-infection risk by ∼80% compared to unvaccinated, previously-infected individuals,^35,36^ and in the United Kingdom hybrid protection relative to unvaccinated, previously-uninfected individuals exceeded 90% for >1 year.^14^ In Sweden, one-and two-dose hybrid protection reduced the pre-Omicron re-infection risk by 58% and 66%, persisting up to 9 months for the latter, compared to unvaccinated, previously-infected individuals.^16^

Perhaps not unexpectedly, we found prior infection improved vaccine protection, and vice-versa, during the Omicron period, consistent with the albeit limited epidemiological evidence elsewhere pertaining to hybrid protection against Omicron^20,34,37^. Among previously-infected HCWs in the United States (US), two-dose mRNA vaccination reduced the risk of symptomatic Omicron re-infection by 64% relative to the once or never-vaccinated (combined grouping), similar to our two-dose estimate of 68% instead relative to the infection-naïve never-vaccinated.^37^ Among previously-infected US adults, the Omicron-hospitalization risk was reduced among the vaccinated vs. unvaccinated by 33%, 35% and 68% with one, two and three mRNA doses^38^. The latter is in contrast to our findings showing that among previously-infected people a third dose did not meaningfully improve the already-substantial two-dose protection against Omicron hospitalization (97% vs. 94%), that was furthermore sustained >1 year.

This study has limitations, notably including that unrecognized/undocumented asymptomatic infections may have led to under-estimation of infection-induced protection. Mathematical modeling, however, suggests such misclassification is likely to have had minimal impact on our findings.^39^ Although we could not control for the bias of differential virus exposure,^40^ similar patterns when directly comparing protected groups (e.g. previously-infected, vaccinated vs. non-infected, vaccinated), are reassuring. Immuno-compromised people were prioritized for an early third dose in Quebec.^12^ To limit under-estimation of three-dose VE associated with their potentially sub-optimal immune responses^41,42^, we excluded specimens from people re-vaccinated at <90-day interval between second and third doses. Our results apply to those surviving their primary infection; with <1% of NAAT-confirmed cases dying, their exclusion will not have meaningfully influenced estimates. Our findings reflect heterologous infection- and/or vaccine-induced protection; homologous protection is anticipated to be higher.^43^ As they were highly correlated, it was not possible to distinguish variation in protection based on VOC-specific vs. time since primary infection.

## Conclusion

Prior heterologous SARS-CoV-2 infection provided substantial and sustained protection against severe outcomes due to Omicron re-infection. Protection was greatest among those previously infected plus vaccinated, whether vaccine came before or after primary infection. In the context of program goals to prevent severe outcomes and preserve healthcare system capacity, our findings suggest that >2 doses of heterologous (ancestral Wuhan-like) vaccine may be of marginal incremental value to previously-infected individuals. Pending vaccine update, such doses may be better prioritized to more vulnerable individuals globally.

## Data Availability

All data produced in the present study are available upon reasonable request to the authors

## Funding

This work was supported by the Ministère de la santé et des services sociaux du Québec.

## Conflict of interest disclosures

All authors have completed the ICMJE Form for Disclosure of Potential Conflicts of Interest. GDS received a grant from Pfizer unrelated to the current work. RG received honoraria as speaker at the RSV workshop financed by Abbvie. JF is chair of the provincial genomic surveillance committee of SARS-CoV-2. Others authors declare no conflict of interest.

## Notes

### Summary of Updates

Correction in supplementary figure 5.

