## Supplemental material for "Protection against Omicron re-infection conferred by prior heterologous SARS-CoV-2 infection, with and without mRNA vaccination"

### Supplementary Material

**Supplementary Figure 1.** Epidemiological curve of reported SARS-CoV-2 cases and vaccination coverage, displayed by month and predominant variant-of-concern periods, Quebec, Canada

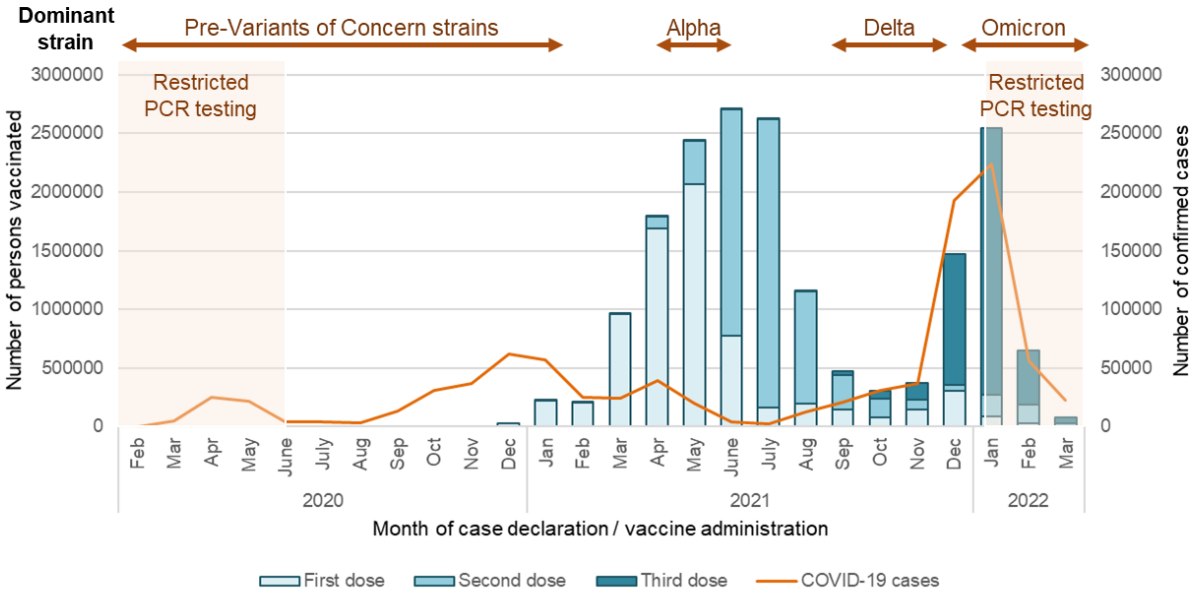

Note: Variant-of-concern (VOC) detection provincially varied over the course of the pandemic in response to changing epidemic patterns, case load and laboratory capacity as well as the profile of emerging and identified VOC. Predominant VOC periods were defined by provincial genomic-based surveillance and were also used to assign likely individual VOC case status.

Before February 2021, VOC detection was undertaken on convenience samples by whole genome sequencing (WGS). From February 1, 2021 to mid-October 2021 screening was systematically undertaken on all specimens. From October 2021 to February 2022 screening was undertaken on a random sample of 5-10% of case viruses, prioritizing travelers, reinfections, outbreaks and severe cases. Between October 9 and December 12, 2021 Delta was almost exclusively identified among characterized viruses. On November 30 and December 14, 2021 all case viruses were screened with Omicron detected in none and 20% of samples, respectively.[1] Conversely, during the first two weeks of the study period after December 25, 2021, 94% and 98% of cases viruses from sentinel laboratories that were genetically screened were Omicron. The BA.2 sub-lineage of Omicron was first detected in Quebec at the end of January 2022, representing 53% of screened specimens by mid-March 2022.[1]

Based on the above, individual VOC status of primary infections occurring before the study period and of cases identified during the study period was assigned as follows:

- Before February 1, 2021: assumed pre-VOC
- February 1 to October 8, 2021: Alpha/Beta/Gamma/Delta as individually diagnosed
- October 9 to December 12, 2021:  assumed Delta
- December 13-25, 2021: excluded based on requirement for ≥90-day delay (see study exclusions)
- December 26, 2021 to March 12, 2022 (study period): assumed Omicron

[1] Institut national de santé publique du Québec. Données sur les variants du SRAS-CoV-2 au Québec. Published April 1, 2022. Accessed April 1, 2022. https://www.inspq.qc.ca/covid-19/donnees/variants

**Supplementary Figure 2.** Exposure categories defined by prior primary SARS-CoV-2 infection and vaccination histories

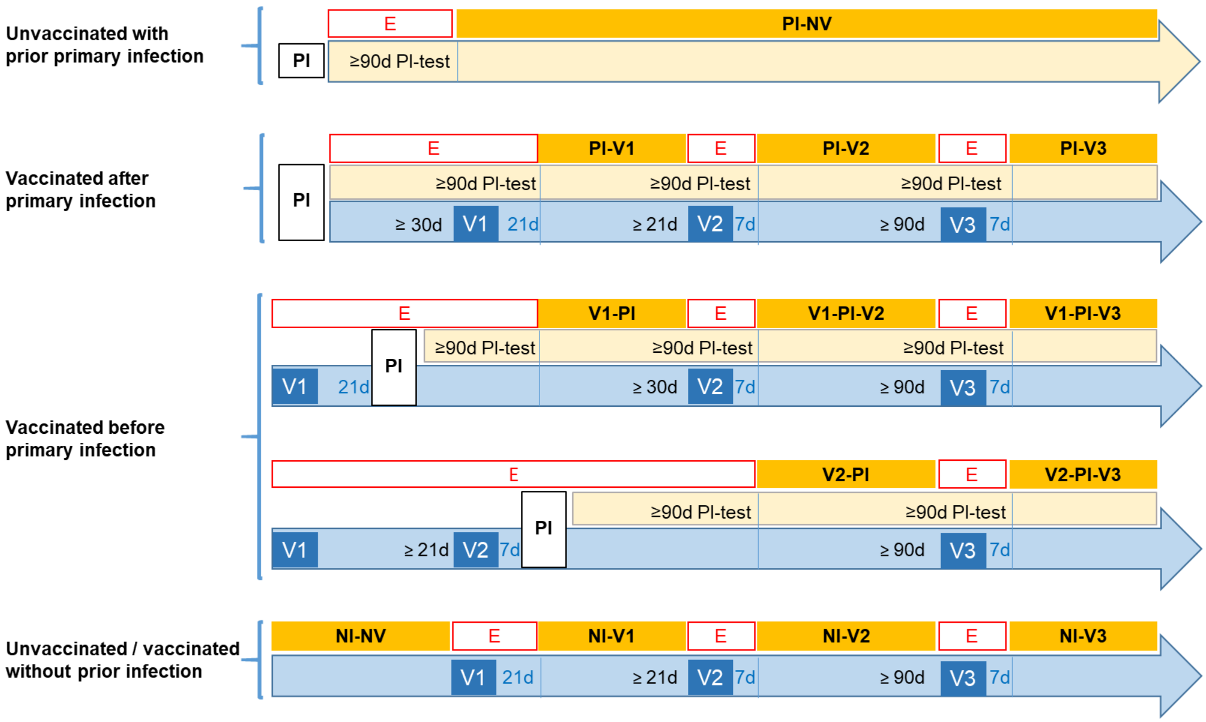

Abbreviations: d, days (between vaccine doses in blue font and between primary infection, testing and vaccine doses in black font); E, exclusion; NI, no infection previously; NI-NV, no prior infection previously non-vaccinated; NI-V1, no infection previously, one vaccine dose; NI-V2, no infection previously, two vaccine doses; NI-V3, no infection previously, three vaccine doses; PI, prior primary infection; PI-NV, prior infection non-vaccinated; PI-V1, prior infection before one vaccine dose; PI-V2, prior infection before two vaccine doses; PI-V3, prior infection before three vaccine doses; V1-PI, prior infection after one vaccine dose; V1-PI-V2, prior infection after first but before second vaccine dose; V1-PI-V3, prior infection after first but before second and third vaccine doses; V2-PI, prior infection after two vaccine doses; V2-PI-V3, prior infection after second but before third vaccine dose; V3-PI, prior infection after three vaccine doses; V1, vaccine dose 1; V2, vaccine dose 2; V3, vaccine dose 3

**Supplementary Figure 3.** Participant flowchart

1,778,623 TAAN tests done between December 26, 2021 and March 12, 2022

1,754,358 (98.6%) tests with unique identifying number correctly merged with immunization registry and clinical case databases

**Exclusions:**
- Long-term care residents;  n = 176,635 (10.1%)
- Reason for testing: confirmation of healing, confirmation of positive antigen test, unknown; n = 156,090 (8.9%)
- Age <12 years old; n = 48,349 (2.8%)
- Reinfection occurring before December 26, 2021; n= 2,402 (0.1%)
- Tests done within 90 days after a primo-infection; n = 29,075 (1.7%)
- Tests done after an initial positive test between December 26, 2021 and March 12, 2022; n = 1,542 (0.1%)
- Vaccination with Astra-Zeneca or Janssen vaccines; n = 19,034 (1.1%)
- Invalid vaccination date; n = 20 (<0.1%)
- Invalid intervals between dose 1 and dose 2 (<21 days); n = 539 (<0.1%)
- Invalid interval between dose 2 and dose 3 (<90 days); n = 9,677 (0.6%)
- Invalid interval between primo-infection and dose 1 (<30 days); n = 3,305 (0.2%)
- Excluded post-vaccination periods (0-20 days post-dose 1, 0-6 days post-dose 2, 0-6 days post-dose 3); n = 52,908 (3.0%)

1,030,775 negative tests (several per person)

224,007 positive tests
9,505 (4.2%) with prior primary infection

472,432 negative tests (1 randomly selected per person)
29,712 (6.3%) with prior primary infection

**Supplementary Figure 4.** Odds ratios for Omicron re-infection by primary SARS-CoV-2 (non-Omicron) infection and/or mRNA vaccination (by number of doses), relative to individuals with no infection history

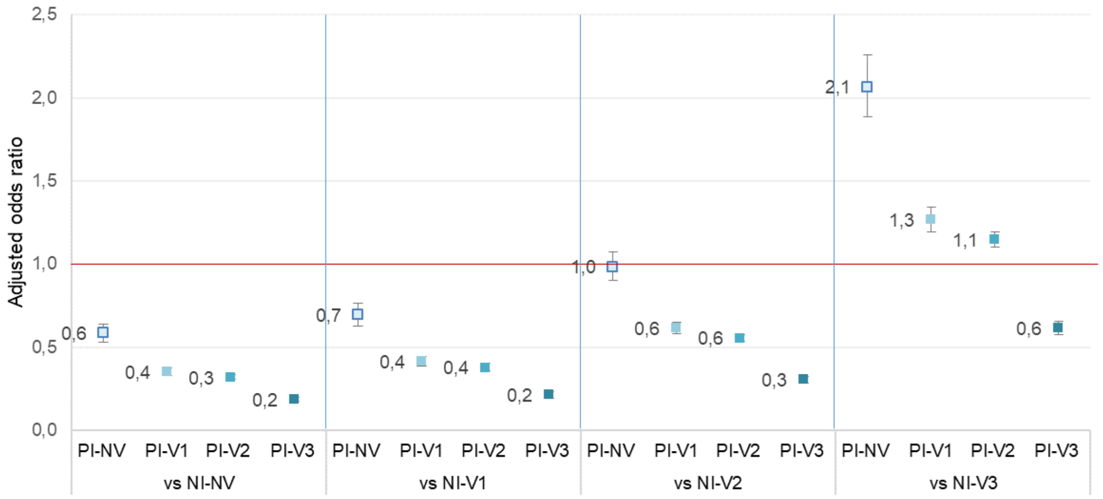

Abbreviations: NI-NV, no prior infection previously non-vaccinated; NI-V1, no infection previously, one vaccine dose; NI-V2, no infection previously, two vaccine doses; NI-V3, no infection previously, three vaccine doses; PI, prior primary infection; PI-NV, prior infection non-vaccinated; PI-V1, prior infection before one vaccine dose; PI-V2, prior infection before two vaccine doses; PI-V3, prior infection before three vaccine doses

Note: Logistic regression models comparing persons with prior primary infection with/without vaccination to those without prior infection with/without vaccination. All estimates adjusted for age (4 categories), sex, indication for testing and epidemiological week

**Supplementary Figure 5.** Odds ratios for Omicron hospitalization by primary SARS-CoV-2 (non-Omicron) infection and/or mRNA vaccination (by number of doses), relative to individuals with no infection history

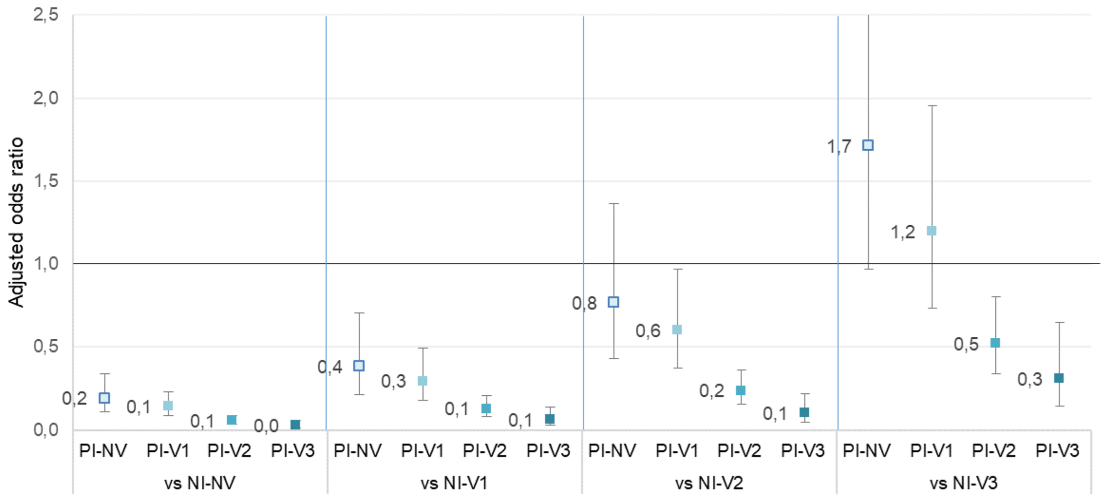

Abbreviations: NI-NV, no prior infection previously non-vaccinated; NI-V1, no infection previously, one vaccine dose; NI-V2, no infection previously, two vaccine doses; NI-V3, no infection previously, three vaccine doses; PI, prior primary infection; PI-NV, prior infection non-vaccinated; PI-V1, prior infection before one vaccine dose; PI-V2, prior infection before two vaccine doses; PI-V3, prior infection before three vaccine doses

Note: Logistic regression models comparing persons with prior primary infection with/without vaccination to those without prior infection with/without vaccination. All estimates adjusted for age (4 categories), sex, indication for testing and epidemiological week
